# Remdesivir for the Treatment of COVID-19: An Updated Systematic Review and Meta-Analysis

**DOI:** 10.1101/2022.01.22.22269545

**Authors:** Todd C. Lee, Srinivas Murthy, Olivier Del Corpo, Julien Senécal, Guillaume Butler-Laporte, Zahra N Sohani, James M. Brophy, Emily G. McDonald

**Author notes:** **Corresponding author:** Dr. Todd C. Lee, Royal Victoria Hospital, 1001 Decarie Blvd Room E5.1820, Montréal, QC Canada H4A3J1, Phone XXX Fax: XXX.

## Abstract

**Background:** The benefits of remdesivir in the treatment of hospitalized patients with Covid-19 remain debated with the National Institutes of Health and the World Health Organization providing contradictory recommendations for and against use.

**Methods:** We performed a systematic review of randomized controlled trials (RCTs) of remdesivir for the treatment of hospitalized patients with COVID-19. The primary outcome was mortality, stratified by oxygen use (none, supplemental oxygen without mechanical ventilation, and mechanical ventilation). We conducted a frequentist random effects meta-analysis on the risk ratio (RR) scale and, to better contextualize the probabilistic benefits, we also performed a bayesian random effects meta-analysis on the risk difference scale.

**Results:** We identified 8 randomized trials, totaling 9157 participants. The RR for mortality comparing remdesivir versus control was 0.71 (95% confidence interval [CI] 0.42-1.22; I^2^=0.0%) in the patients who did not require supplemental oxygen; 0.83 (95%CI 0.73-0.95; I^2^=0.0%) for nonventilated patients requiring oxygen; and 1.19 (95%CI 0.98-1.44 I^2^=0.0%) in the setting of mechanical ventilation. Using neutral priors, the probabilities that remdesivir reduces mortality were 74.7%, 96.9% and 8.9%, respectively. The probability that remdesivir reduced mortality by more than 1% was 88.1% for nonventilated patients requiring oxygen.

**Conclusion:** Based on this meta-analysis, there is a high probability that remdesivir reduces mortality for nonventilated patients with COVID-19 requiring supplemental oxygen therapy.

## Introduction

The World Health Organization (WHO) recommends against the use of remdesivir^1^ for all patients with Covid-19, based on the results of the SOLIDARITY trial, which failed to demonstrate a reduction in hospital length of stay or mortality^2^. By contrast, guidelines from the National Institutes of Health^3^ and the Infectious Diseases Society of America^4^ recommend the remdesivir in the treatment of Covid-19 for patients who do not require mechanical ventilation. These recommendations follow the completion of the Adaptive Covid-19 Treatment Trial 1 (ACTT-1),^5^ which demonstrated a substantial decrease in hospital length of stay. On an international level, the benefits of remdesivir for the treatment of Covid-19 therefore remain debated and, in many countries, treatment with remdesivir may be underutilized.

We previously hypothesized that conflicting trial results relate to the differential effects of remdesivir as a function of the severity of the underlying illness. We tested this hypothesis in January 2021, when we conducted a bayesian meta-analysis to determine the probability that remdesivir reduces mortality as a function of oxygen requirements^6^. Our findings suggested that the probability of any mortality benefit was 69% among patients without oxygen requirements, 92% in those requiring supplemental oxygen who were not ventilated, and only 7% among patients requiring mechanical ventilation. Since this time, two large new trials comparing remdesivir versus standard of care have been published. We therefore conducted a systematic review and meta-analysis to clarify whether remdesivir reduces mortality in moderately ill hospitalized patients with Covid-19.

## Methods

### Search Strategy, Study selection, and Data Extraction

We searched PubMed from January 1^st^, 2020, to January 21, 2022 in order to identify randomized controlled trials comparing remdesivir to placebo or standard of care. New trials were added to our previous results^6^. We used the search syntax “remdesivir AND (randomized OR randomised) AND 2021-01-15[dp]:2022-01-21[dp]”. Two independent reviewers screened for eligibility. The trial inclusion criteria and patient demographics are summarized in **Table 1** for included studies. Two reviewers independently extracted the outcome of all-cause mortality stratified by level of baseline oxygen support. Oxygen support was defined according to categories in the largest trial, SOLIDARITY, as: (i) no oxygen required; (ii) supplemental oxygen (without mechanical ventilation); and (iii) mechanical ventilation.

**Table 1.**
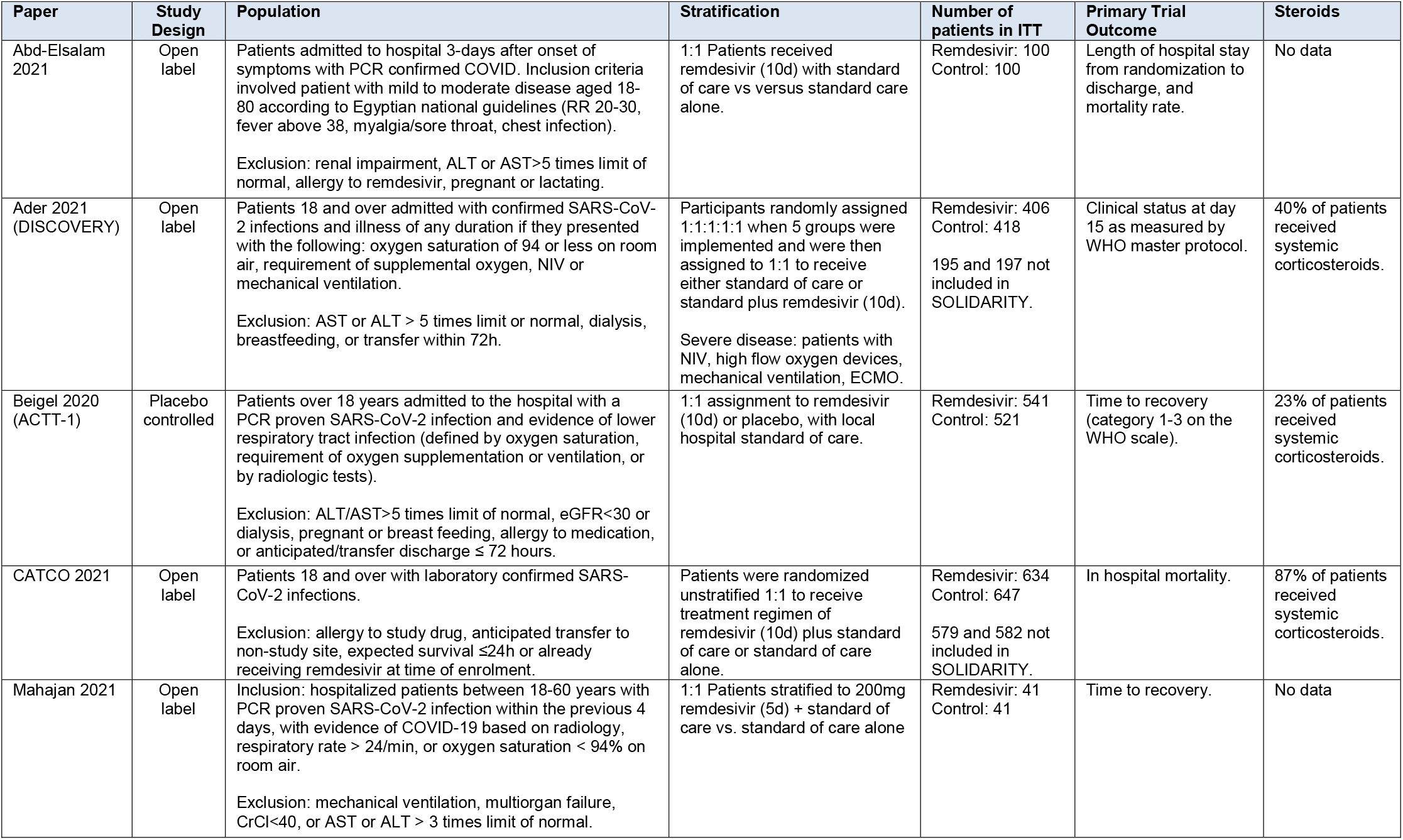
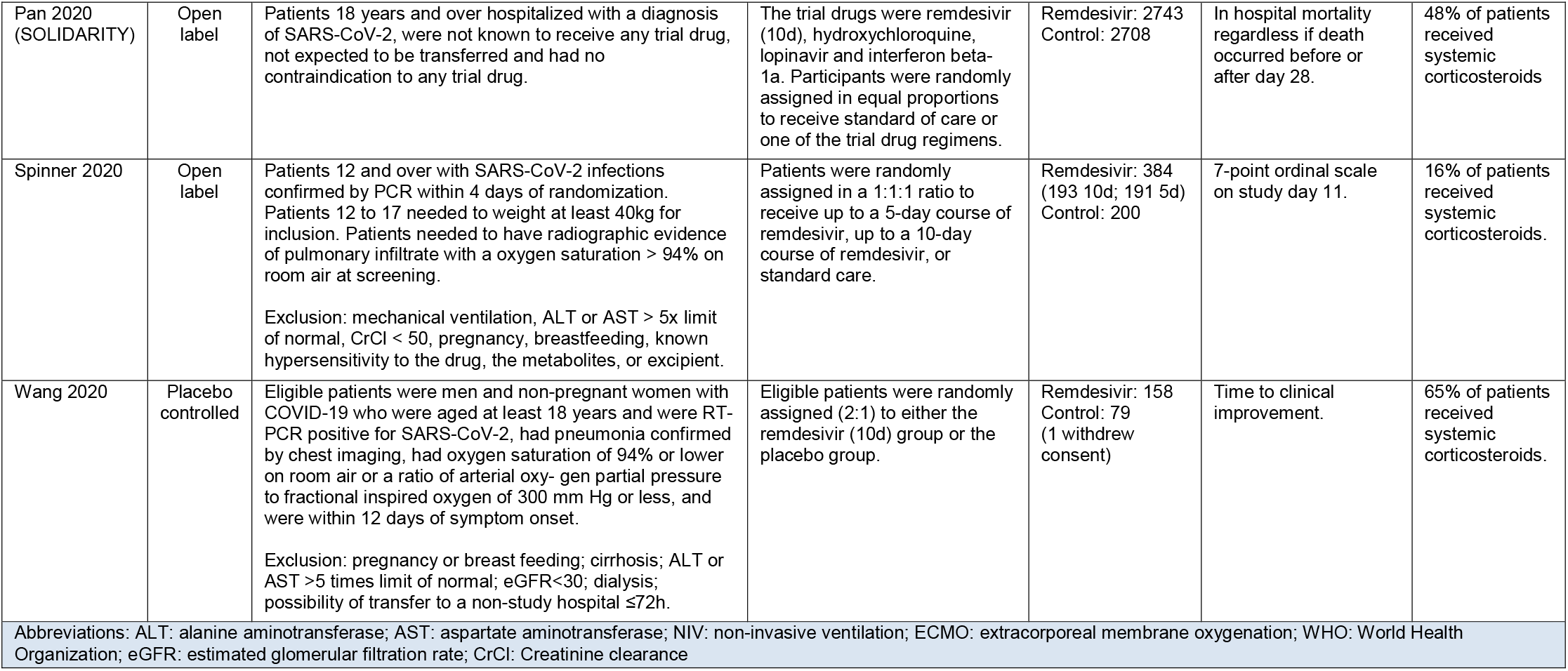
Description of Included Trials.

### Assessment of Bias

Two independent reviewers assessed each study for bias using the Cochrane risk-of-bias 2 tool for randomized trials.

### Meta-Analysis

Study results are reported according to the PRISMA (Preferred Reporting Items for Systematic Reviews and Meta-analyses) 2020 checklist^7^. We conducted both a frequentist and a bayesian meta-analysis both stratified by the level of oxygen support. A DerSimonian–Laird random effects meta-analysis on the risk ratio (RR) scale was used to undertake our frequentist analysis using the metan^8^ command in STATA version 17 (STATACorp, USA). Next, to quantify the mortality benefit in absolute terms and to address clinically meaningful differences (a priori defined as the probability of achieving at least a 1% absolute mortality reduction), we conducted a bayesian meta-analysis on the risk difference scale using R^9^ and the bayesmeta package^10^. Vague proper non-informative priors were used: μ centered at 0 (standard deviation = 4), which corresponds to no effect; and heterogeneity τ assumed to be half-normal prior with a scale of 0.03^6^. Figures of posterior density vs. absolute differences in mortality between remdesivir and control patients were generated, and we integrated the area under the curve to obtain the probability for any mortality benefit and for a benefit exceeding 1% respectively.

### Certainty Assessment

Certainty of evidence for hospitalization was assessed using the grading of recommendations assessment, development, and evaluation (GRADE) approach^11^. Two reviewers with familiarity and experience with GRADE rated each domain separately; discrepancies were resolved by consensus. Certainty was rated as high, moderate, low, or very low, based on the GRADE domains.

## Results

Search results in January 2021 included 4 trials; the present search yielded an additional 148 articles, of which 5 new trials were reviewed for eligibility for inclusion^12–16^ (**Supplemental Figure 1)**. One of the trials only contained patients previously reported in SOLIDARITY and was thus excluded^13^. A total of 8 RCTs were included in the present analysis.

The DISCOVERY and CATCO trials were previously partially reported as part of the SOLIDARITY trial; therefore, to avoid duplication, we obtained data directly from the study teams on the subset of patients who were not already included in SOLIDARITY.

Results of the Mahajan et al.^15^ study were not presented as intention-to-treat. We therefore reanalyzed their data using the intention-to-treat principle. We also included participants who were discharged before day 12 (categorized as alive), as well as those who died before day 12 (categorized as deceased).

Some trials deviated from the oxygen support categories described in the SOLIDARITY trial. We made the following adjustments to include them in our analyses. For the trial by Wang et al.^17^, although study inclusion criteria required the use of oxygen, 3 patients in the placebo group were not receiving oxygen at the time of their first dose of remdesivir. Further, there was one mechanically ventilated patient in the placebo group. We included results of this trial in the ‘supplemental oxygen without mechanical ventilation’ group. For the trial by Spinner et al.^18^, although oxygen requirement was a study exclusion criterion, 14% and 19% of remdesivir and control patients, respectively, developed a need for supplemental oxygen between screening and the first dose remdesivir. However, results were not reported by day 1 oxygen requirements. As most patients did not require supplemental oxygen, and due to the overall low mortality rate in both arms, we included this study in the ‘no oxygen support’ group. Next, the participants in the DISCOVERY trial^12^ were classified according to disease severity. Moderate disease severity (no oxygen [16 total patients] and oxygen by nasal prongs or mask) and severe disease (high flow nasal oxygen, non-invasive, and invasive ventilation). We assigned the moderate group (n=223) to ‘supplemental oxygen without mechanical ventilation’ and the severe group (n=169) to ‘mechanical ventilation’. Finally, the trial by Abd-Elsalam et al.^14^ included mild and moderate severity patients with an average oxygen saturation of 87% and 89% in the remdesivir and control groups respectively. Although this study did not report results stratified by baseline oxygen requirements, mechanical ventilation was a trial exclusion criterion. We assigned these patients to the ‘supplemental oxygen without mechanical ventilation’ subgroup.

### Included studies

The meta-analysis includes 8 trials (**Table 1**)^2,5,12,14–18^ and 9157 unique patients (2148 without oxygen, 5974 nonventilated with supplemental oxygen, and 1035 mechanical ventilation; **Figure 1**). We included all randomized controlled trials comparing remdesivir to placebo or standard of care that recruited symptomatic inpatients with microbiologically confirmed SARS-CoV-2 infection. All studies were considered at overall low risk for bias (**Supplemental Figure 2)** although there were some concerns with the two smaller independent studies^14,15^. While 6 of 8 studies were not placebo controlled, we believed there was low risk of bias considering the outcome of all-cause mortality.

**Figure 1.**
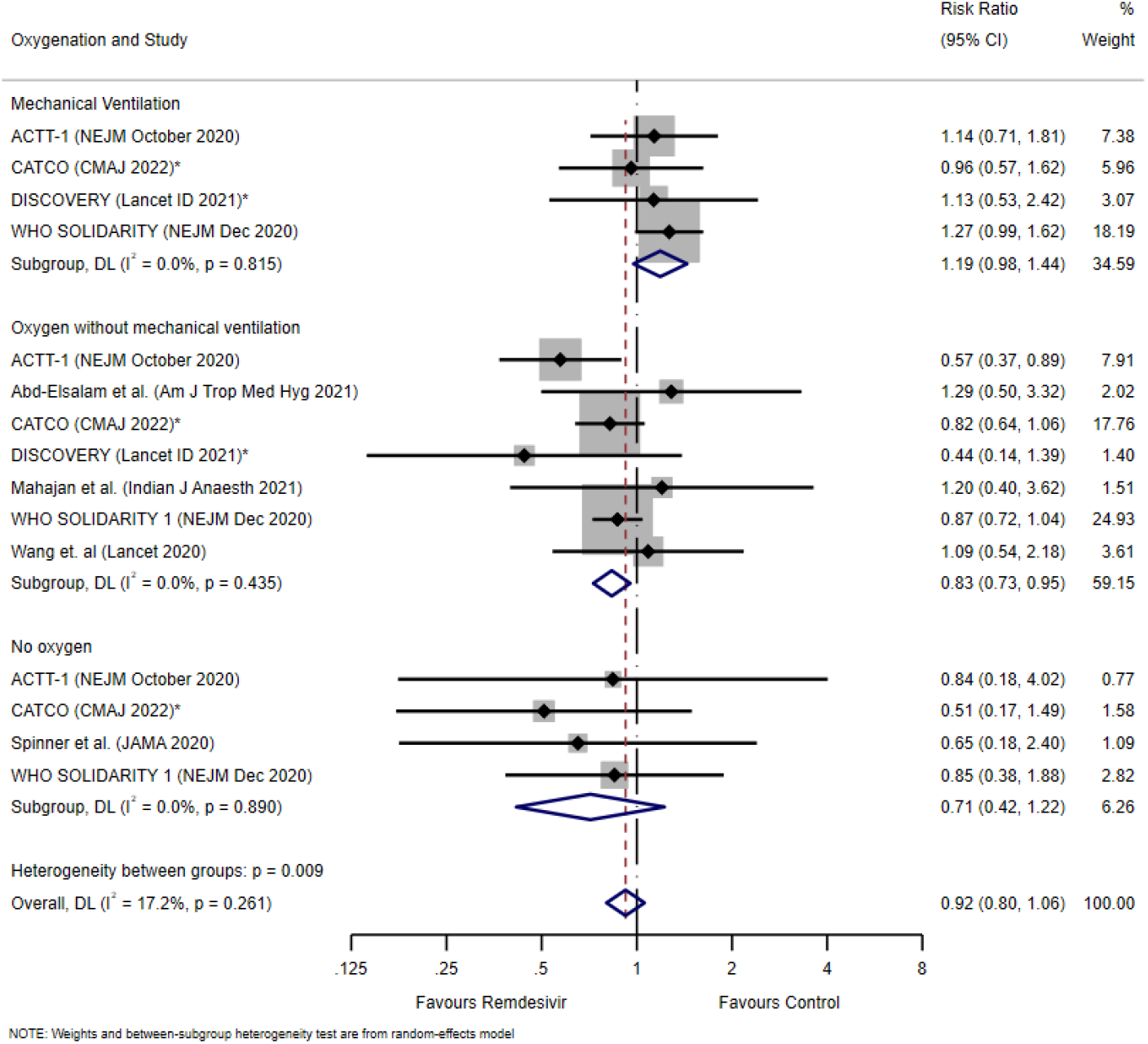
Random Effects Meta-Analysis. *Excludes patients already included in SOLIDARITY (NEJM 2020)

### Meta-analysis

With respect to the primary outcome of mortality, treatment with remdesivir was associated with a RR and 95% confidence interval (CI) of 0.71 (95%CI 0.42-1.22; I^2^=0.0%) for patients without oxygen; 0.83 (95%CI 0.73-0.95; I^2^=0.0%) for patients requiring oxygen, and 1.19 (95%CI 0.98-1.44; I^2^=0.0%) for those on mechanical ventilation (**Figure 1)**. For patients without oxygen the probability of any mortality benefit was 74.7%, for those requiring oxygen 96.9%, and for those on mechanical ventilation was 8.9% (**Figure 2**). In particular, for patients requiring oxygen but not mechanical ventilation, the mean estimate for absolute risk difference was 2.4% and the probability that the absolute risk reduction was greater than 1% was 88.1%.

**Figure 2.**
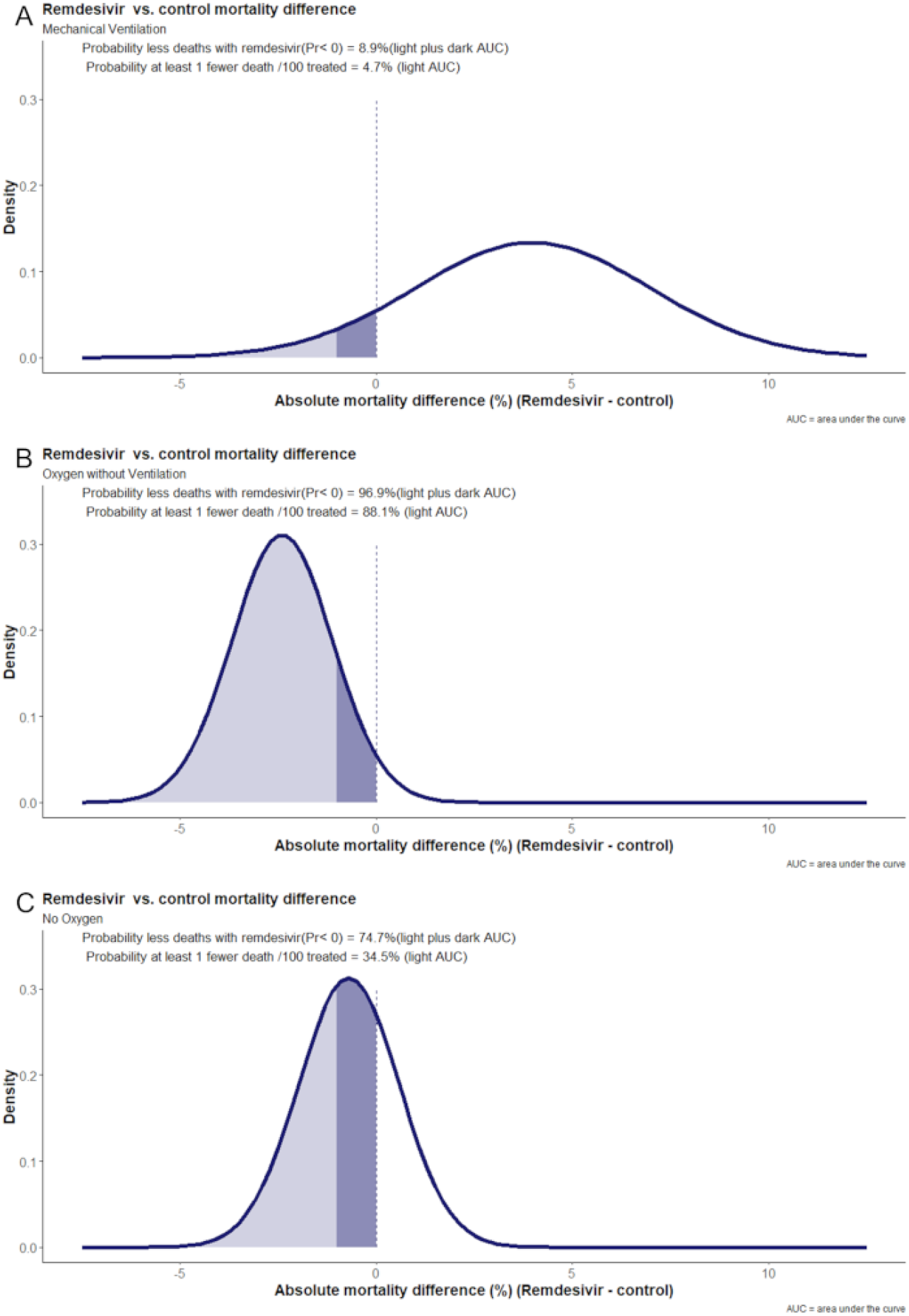
Probability density functions for combined posterior distributions of the included remdesivir trials. (A) Mechanical ventilation. (B) Supplemental oxygen without mechanical ventilation. (C) No oxygen support.

### GRADE Certainty of Evidence

Regarding the overall certainty of the evidence, the primary outcome of our analysis was mortality, which is not likely subject to adjudication bias. However, most of the included studies were open label, and there was also the potential for misclassification of oxygen requirements, reducing the overall certainty of the evidence away from high. The probability of benefit in the oxygenated subgroup, and correspondingly of harm in the mechanical ventilation subgroup, were both high, in addition to being consistent with our prior analyses. In these respective subgroups, a recommendation for and against remdesivir is proposed with moderate certainty. It should be noted that participants requiring high flow nasal cannula and non-invasive ventilation were underrepresented in the included trials rendering the certainty of evidence weak for this subgroup. Finally, the suggestion of mortality benefit in patients who do not require oxygen is of weak certainty, given the probability of a meaningful effect was very modest.

## Discussion

Our meta-analysis comparing remdesivir versus placebo or standard of care suggests a high probability of a clinically meaningful reduction in mortality for patients requiring supplemental oxygen. Although an analysis of remdesivir trials stratified by oxygen requirements is *post hoc*, the ACTT-1 trial^5^ already suggested a potential mortality benefit for patients in the “Goldilocks zone” (disease severity requiring oxygen without needing critical care). By contrast, we found a high probability that remdesivir harms patients requiring mechanical ventilation and that any beneficial effect size is much smaller for patients who did not require any supplemental oxygen.

There are still unanswered questions related to remdesivir treatment in hospitalized patient subgroups, which could be the focus of future randomized trials. For example, whether there is a benefit in early nosocomial Covid-19, or “incidental” non-hypoxemic Covid-19 in patients high risk for deterioration. This could be akin to the benefit observed in the recent PINETREE trial that demonstrated superiority of 3 days of remdesivir versus placebo in high risk outpatients^19^. Likewise, the role of remdesivir in the setting of high flow nasal oxygen or non-invasive ventilation needs to be clarified as, to date, this population is less represented in trials, or the total data is not sufficiently granular.

There are limitations to this analysis, the principal one being that the standard of care for Covid-19 continues to evolve at a staggering pace. Earlier in the pandemic, trial participants were less likely to receive treatments now known to reduce adverse outcomes including steroids, monoclonal antibodies, immunomodulatory therapies, or therapeutic anticoagulation.

Additionally, very few of the participants included in this analysis were vaccinated against Covid-19 and all results predate the delta and omicron variants. Whether there will be additional large randomized controlled trials of remdesivir in vaccinated patients with newer variants remains to be seen and so inferring a precise magnitude of benefit of remdesivir in these populations is challenging. A final limitation we wish to note is a small lack of granularity with respect to oxygen requirements for a handful of patients; in this case, an individual patient meta-analysis could provide more precise results and transparent data reporting and sharing is welcomed. The strengths of this analysis are the avoidance of duplicated patients despite the inclusion of published SOLIDARITY country-level studies, our *a priori* decision to stratify the analysis by oxygen requirements, and the added bayesian analysis to contextualize the probability of a reduction in mortality from remdesivir.

## Conclusions

There is a high probability (97%) that remdesivir reduces mortality for patients who require oxygen but who are not yet critically ill. Future antiviral treatment trials for noncritically ill hospitalized patients with Covid-19 should likely include remdesivir as an active treatment arm, stratified by oxygen requirements. Importantly, we hope the results of this meta-analysis support harmonization of discrepant international guideline recommendations and facilitate the appropriate uptake of remdesivir in certain patient populations.

## Supporting information

PRISMA Checklist

## Data Availability

All data produced in the present work are contained in the manuscript.

## CRediT Author Statement

The first author named is the lead author and corresponding author. The last author is the senior author. We describe contributions to the paper as follows:

*Conceptualization* – TCL and EGM; *Methodology* – TCL, and JMB; *Validation* – TCL; *Formal analysis* – TCL and JMB; *Data Curation –* TCL, SM, ODC, JS, and GBL; *Writing – Original Draft* – TCL *Writing – Review and Editing* – all listed; *Visualization* – TCL, JMB and EGM; *Supervision* – TCL.

## Ethical approval

Not required.

## Registration

As this was an update of previous work based on newly published information, this systematic review was not pre-registered, and a specific protocol was not prepared.

## Data Sharing

All data required to replicate this analysis is included in the manuscript. Statistical code is available for audit upon written request.

## Acknowledgements

The authors wish to thank all the DISCOVERY trial team and the CATCO team for providing the unpublished data required to complete this analysis.

