## Supplementary Figures for "Remdesivir for the Treatment of COVID-19: An Updated Systematic Review and Meta-Analysis"

**Supplemental Figure 1 – PRISMA diagram**

**Identification of studies via Database**

**databases and registers**

Records removed *before screening*:

None

Records identified from:

PubMed (n=148)

**Identification**

Records screened

(n=148)

Records excluded

Not RCTs (n = 142)

Outpatient RCT (n=1)

Reports not retrieved

None

Reports sought for retrieval

(n =5)

**Screening**

Excluded:

All patients already published (n=1)

Reports assessed for eligibility

(n=5)

Studies included in review

(n =4)

Reports of included studies

(n =4)

**Included**

*From:*  Page MJ, McKenzie JE, Bossuyt PM, Boutron I, Hoffmann TC, Mulrow CD, et al. The PRISMA 2020 statement: an updated guideline for reporting systematic reviews. BMJ 2021;372:n71. doi: 10.1136/bmj.n71

**Supplemental Figure 2 – Risk of Bias Assessment**

|  | Random sequence generation | Allocation concealment | Blinding of participants and personnel | Blinding of outcome assessment | | Incomplete outcome data | | Selection reporting | Other bias |
| --- | --- | --- | --- | --- | --- | --- | --- | --- | --- |
| Abd-Elsalam 2021 | Low | Low | Low | Low | | Some concerns | | High | Low |
| ACTT-1 2020 | Low | Low | Low | Low | | Low | | Low | Low |
| CATCO 2021 | Low | Low | Low | Low | | Low | | Low | Low |
| DISCOVERY 2021 | Low | Low | Low | Low | | Low | | Low | Low |
| Mahajan 2021 | Low | Some concerns | Low | Low | | Some concerns | | High | Low |
| Spinner 2020 | Low | Low | Low | Low | | Low | | Low | Low |
| Wang 2020 | Low | Low | Low | Low | | Low | | Low | Low |
| WHO SOLIDARITY 2020 | Low | Low | Low | Low | | Low | | Low | Low |
